# Preventing a cluster from becoming a new wave in settings with zero community COVID-19 cases

**DOI:** 10.1101/2020.12.21.20248595

**Authors:** Romesh G Abeysuriya, Dominic Delport, Robyn M Stuart, Rachel Sacks-Davis, Cliff C Kerr, Dina Mistry, Daniel J Klein, Margaret Hellard, Nick Scott

**Affiliations:** Burnet Institute, Melbourne, Victoria, Australia; Department of Epidemiology and Preventive Medicine, Monash University; Department of Mathematical Sciences, University of Copenhagen, Copenhagen, Denmark; Institute for Disease Modeling at the Bill & Melinda Gates Foundation, Seattle, USA; School of Physics, University of Sydney, Sydney, New South Wales, Australia

## Abstract

In settings with zero community transmission, any new SARS-CoV-2 outbreaks are likely to be the result of random incursions. The level of restrictions in place at the time of the incursion is likely to considerably affect possible outbreak trajectories. We used an agent-based model to investigate the relationship between ongoing restrictions and behavioural factors, and the probability of an incursion causing an outbreak and the resulting growth rate. We applied our model to the state of Victoria, Australia, which has reached zero community transmission as of November 2020.

We found that a future incursion has a 45% probability of causing an outbreak (defined as a 7-day average of >5 new cases per day within 60 days) if no restrictions were in place, decreasing to 23% with a mandatory masks policy, density restrictions on venues such as restaurants, and if employees worked from home where possible. A drop in community symptomatic testing rates was associated with up to a 10-percentage point increase in outbreak probability, highlighting the importance of maintaining high testing rates as part of a suppression strategy.

Because the chance of an incursion occurring is closely related to border controls, outbreak risk management strategies require an integrated approaching spanning border controls, ongoing restrictions, and plans for response. Each individual restriction or control strategy reduces the risk of an outbreak. They can be traded off against each other, but if too many are removed there is a danger of accumulating an unsafe level of risk. The outbreak probabilities estimated in this study are of particular relevance in assessing the downstream risks associated with increased international travel.

## Introduction

Strategies to contain SARS-COV-2 vary from setting to setting, depending on epidemiological factors, health system capacity, political will, and the economic feasibility of introducing or maintaining physical distancing restrictions. In settings with high prevalence, policy decisions are heavily guided by epidemic indicators such as the number of diagnoses per day, or the number of people hospitalized and the health system capacity. Policy changes affect these indicators within weeks, so these measures provide feedback to guide scaling interventions up or down. However, in settings such as New Zealand and Taiwan, that have effectively eliminated community transmission of the virus as of December 2020, the situation is different. With zero community transmission, epidemic indicators are not available to guide policy, and the risk to the community is difficult to quantify since even large, high-mixing events will not lead to an outbreak. Nonetheless, a level of risk persists due to the possibility that an external source can seed an undiagnosed infection and lead to an outbreak. This may have important consequences if the outbreak grows too rapidly or is detected too late to be contained with a combination of testing, contact tracing and quarantine. Such an outbreak would likely occur without any advance warning– for example, an individual who has returned from overseas and was quarantined, but remained undetected but infectious after the prescribed quarantine period (usually 14 days), or a failure in quarantine procedure leading to a transmission event to support staff during quarantine. These kinds of events have now been observed on several occasions in low prevalence settings, including recent Australian outbreaks in Melbourne, Adelaide, and Sydney, with the Melbourne outbreak leading to considerable restrictions on movement and social mixing that lasted almost six months.

If an undetected infection is introduced to a setting with no community transmission, three broad outcomes are possible: no/limited transmission occurs, and the outbreak dissipates naturally (perhaps even without being diagnosed and detected); transmission occurs leading to a cluster of infections that is eventually contained with testing, contact tracing and quarantine; or sufficient transmission occurs to create sustained epidemic growth such that additional restrictions or interventions are required to regain epidemic control. Which outcome will occur is stochastic, and depends on a number of factors including the extent of restrictions imposed at the time (and hence the extent of individual-level mixing that is allowed), the capacity and efficiency of the testing and contact tracing system, compliance with quarantine directions, any non-pharmaceutical interventions in place at the time (e.g. physical distancing policies, mask coverage), and the socio-demographic and contact networks of the first few infected individuals.

A range of outbreak outcomes have been seen in various settings over the second half of 2020. In Australia, following an initial wave of SARS-CoV-2 infections in March-April 2020, a variety of restrictions and public health measures were imposed to reduce transmission, and by May 2020 all Australian jurisdictions had negligible community transmission. After easing restrictions, in late June the state of Victoria experienced an epidemic resurgence requiring restrictions to be re-imposed between July and August, with a second wave peak of 687 diagnoses on 4th August (1). Almost all cases were able to be traced back to just four incursion events. Over the same period, the states of NSW and Queensland also detected instances of community transmission. However, in these states the outbreak was controlled or contained with testing and contact tracing, without requiring major restrictions to be imposed. Similarly, New Zealand experienced several incursion events after reaching zero community transmission. In August 2020, a single infection triggered a three-week lockdown in Auckland, with 179 downstream infections in total (2), but subsequent incursions in October and November did not spread widely and were contained without community restrictions. These different examples highlight how a range of different outcomes are possible when new cases are seeded, even under similar circumstances.

Following the late-2020 outbreak in Victoria, the state has now achieved zero detected community transmission (1). As it moves into this transmission regime, decisions need to be made on which restrictions or interventions should be maintained longer-term to balance the competing needs of minimizing outbreak risk and maximizing social and economic freedoms. Maintaining heavier restrictions is likely to significantly reduce the risk of an outbreak if a new case is introduced but would have unsustainable social and economic costs. On the other hand, with no restrictions in place, high levels of mixing and possible relaxed behaviours combined with likely low testing rates due to reduced perceived likelihood of virus transmission, it is possible that a newly introduced case could go unnoticed for weeks, and hence that the introduction of a single case could lead to an uncontrolled outbreak. A happy medium may be a set of lighter restrictions (e.g. limits on large events, mandatory masks in places such as public transport and enclosed public spaces) that have a lower social and economic cost but could still reduce outbreak risk in a meaningful way. To inform what might be appropriate, evidence is needed to quantify the outbreak risks associated with different restrictions and policies, so that they can be weighed against the social and economic cost in an objective way.

In this study we use an agent-based model, Covasim, to quantify the outbreak risks associated with a range of realistic restrictions. For an undetected infection being introduced to a setting with no community transmission, we aimed to estimate the risk of an outbreak and how this risk could be reduced with behavioural changes. The modelling is undertaken using parameters for the state of Victoria, Australia, based on a context of no community transmission.

## Methods

### Model overview

We used an established agent-based microsimulation model, Covasim (3, 4), developed by the Institute for Disease Modeling (USA) and previously adapted by the Burnet Institute to model the Victorian epidemic (5). In brief, agents in the model are assigned an age (which affects their disease prognosis), a household, a school (for people age 5-18) or a workplace (for people aged 18-65), and can participate in a number of daily community activities including attending restaurants, pubs, places of worship, community sport, and small social gatherings. Full details of included contact types, transmission probabilities, and contact tracing capability are provided in the supplement. The model also includes testing, contact tracing and quarantine of close contacts, isolation of confirmed cases, masks, density limits in venues (e.g. 1 person per 4 square metres in restaurants), and other policy restrictions to prevent or reduce transmission in different settings (e.g. closing schools or venues).

### Testing, contact tracing and quarantine

Testing was modelled such that 50% of people with symptoms would seek testing, with a delay of 24 hours between first symptoms and test-seeking. We assumed positive test results take 24 hours to become available (6) and we assumed 75% compliance with self-quarantine while waiting for test results. Following a positive test, all household contacts in the model are notified by the confirmed case directly. Contact tracing for other contacts was assumed to take an additional 24 hours (based on estimated average performance; see (7)), and we assumed it is able to identify 95% of people in workplaces, schools, childcare, and aged care; 50% in of people in venues such as restaurants, and 10% of people in community settings such as public transport. In the model, contact tracing for non-household contacts can be performed for a maximum of 250 newly diagnosed people per day, which is an estimate of Victoria’s tracing capacity during the second wave. Accounting for tracing capacity is important in large outbreaks, but we note that the analyses in this study focus on outbreaks that are much smaller than the tracing capacity, and thus our findings largely do not depend on this parameter. People identified as close contacts of a confirmed case are required to self-quarantine for 14 days, and we assume full compliance with this requirement. It was assumed that 90% of people with symptoms in quarantine would seek testing without delay. A full listing of tracing parameters by layer is provided in the Supplementary Material.

### Masks

A comprehensive meta-analysis (8) (conducted after two others (9, 10)), covering 41 studies of mask effectiveness concluded that that masks are associated with a reduction in infection for mask-wearers by a third compared to control groups. However, mask usage varies with settings, so we classified each contact layer in the model as having high, medium, or low usage. We therefore assumed masks would provide a reduction in transmission probability per contact of 30% in workplaces, entertainment venues, large events, and aged care; 25% in community settings, places of worship, public parks, social gatherings and on public transport; and 10% in cafes, restaurants, pubs and bars.

### Restriction levels

We defined a representative range of restriction levels from normal activity (Level 0) to the hard lockdown (Level 9), as shown in Table 1. These representative levels were based on the staged restrictions used in Victoria, but map approximately to restriction levels in many settings, such as the four-level alert system in New Zealand (11) or the three-tier system in the United Kingdom (12). Full details of each policy and the effect on transmission in each contact layer are provided in the supplement.

**Table 1.** Combinations of policies included in each policy package examined. The specific effects of each policy on disease transmission is provided in the Supplementary Material.

| Policy | Level 0 | Level 1 | Level 2 | Level 3 | Level 4 | Level 5 | Level 6 | Level 7 | Level 8 | Level 9 |
| --- | --- | --- | --- | --- | --- | --- | --- | --- | --- | --- |
| Outdoor gatherings | Unrestricted | Unrestricted | Unrestricted | <50 | <50 | <10 | <10 | <2 people | <2 people | <2 people |
| Small social gatherings | Allowed | Allowed | Allowed | Allowed | Allowed | Allowed | Allowed | Banned | Banned | Banned |
| Childcare | Open | Open | Open | Open | Open | Open | Open | Open | Open | Closed |
| Schools | Open | Open | Open | Open | Open | Open | Open | Open | Primary + 11/12 | Closed |
| Work | Unrestricted | Work from home | Work from home | Work from home | Work from home | Work from home | Some restrictions | Some restrictions | Some restrictions | Heavy restrictions |
| Community sports | Allowed | Allowed | Allowed | Allowed | Allowed | Cancelled | Cancelled | Cancelled | Cancelled | Cancelled |
| Places of worship | Open | Open | Open | 4sqm | Closed | Closed | Closed | Closed | Closed | Closed |
| Cafes/restaurants | Open | Open | Open | 4sqm | Take-away | Take-away | Take-away | Take-away | Take-away | Take-away |
| Pubs/bars | Open | Open | Open | 4sqm | Closed | Closed | Closed | Closed | Closed | Closed |
| Masks | No | No | Yes | Yes | Yes | Yes | Yes | Yes | Yes | Yes |
| Entertainment venues | Open | Open | Open | Open | Closed | Closed | Closed | Closed | Closed | Closed |
| Mobility restrictions | No | No | No | No | No | No | No | No | No | Yes |

### Model calibration

The model was calibrated to the outbreak in Victoria over the June-August period (1), and the associated policy changes and interventions that were implemented over that period. Even though this analysis is based around the introduction of an infection in the context of no community cases, this calibration was used as a method for estimating and validating model parameters for the transmission probability per contact per day in a variety of settings, and the effectiveness of interventions.

### Outbreak analysis

We investigated the probability that a new infection would cause an outbreak if it were introduced to a community setting that had no existing infections. A 60-day period was modelled following the introduction of a single new infection under each restriction level, to represent the period at the start of an outbreak where containment would be managed by ongoing testing, tracing, and isolating procedures rather than being dominated by policy responses (e.g. wider lockdowns and closures prompted by the outbreak, which are uncertain and will vary from setting to setting).

When a new infection is introduced in the model, the epidemic trajectory is random and depends on factors such as who is initially infected (e.g., whether they have many workplace or community contacts, size of their household, or their individual transmissibility) and how quickly they are diagnosed. Therefore 1000 simulations were run for each restriction level, with different initial infections each time.

The are many possible ways to define and classify outbreaks based on metrics such as their size or growth rate. For the purposes of this analysis we have classified simulations as either *‘contained’* (the 7-day average of new cases per day was 0 after 60 days), *‘under control’* (the 7-day average of new cases per day was >0 after 60 days, but did not exceed 5 during the simulation), or an *‘outbreak’* (the 7-day average of new cases per day reached >5 within 60 days). These definitions were chosen based on whether a policy change or additional restrictions/intervention would likely be required. Note that the definitions are in terms of number of diagnoses rather than raw number of infections, because this is a measurable quantity that could form the basis for potential policy responses.

For each simulation it was recorded: (a) whether the infection was contained, under control, or led to an outbreak (as defined above); (b) days to first diagnosis; (c) cumulative number of infections at the time of first diagnosis; (d) days to reach a 7-day average of 5 cases per day; and (d) cumulative number of infections after 60 days.

### Sensitivity analysis: risk reduction strategies

To examine the scope for behavioural changes to complement restrictions, and the risks associated with complacency, we performed sensitivity analyses around the proportion of people with symptoms that seek testing, test-related delays, and test quarantine compliance, as shown in Table 2. Test related delays include delay to test and test turnaround time. The ‘delay to test’ is the time taken between a person becoming symptomatic, and seeking a test, if they get tested. The ‘test turnaround time’ is the time taken between the test being performed and the results becoming available. In Victoria, individuals must self-quarantine if they have been tested and are awaiting results. Thus, they do not quarantine during their delay to test, but do quarantine) during the test turnaround time. The ‘test quarantine compliance’ is the proportion of people that comply with this requirement and quarantine until they receive their test results.

**Table 2.** Parameter scenarios examined for each restriction level.

|  | Scenario | Symptomatic test proportion | Delay to test (days) | Test turnaround time (days) | Test quarantine compliance |
| --- | --- | --- | --- | --- | --- |
| Better | Best case | 0.75 | 1 | 1 | 1 |
|  | More testing | 0.75 |  |  |  |
|  | More test quarantine compliance |  |  |  | 1 |
|  | **Baseline** | **0.5** | **1** | **1** | **0.75** |
| Worse | Less test quarantine compliance |  |  |  | 0.5 |
|  | Slower test results |  |  | 2 |  |
|  | Slower to seek testing |  | 2 |  |  |
|  | Less testing | 0.25 |  |  |  |

## Results

### Outbreak probability

For all restriction levels, a newly seeded undiagnosed case was contained in at least 50% of simulations (Figure 1), but the chance of containment decreased if fewer restrictions were in place. As restrictions were eased, the likelihood of the situation not being contained but either “under-control” or an “outbreak” increased. With no restrictions, 45% of simulations resulted in an outbreak, compared to 40% if working from home was in place (Level 1), 28% with working from home + masks (Level 2), or 23% with working from home + masks + density limits on venues (Level 3).

**Figure 1.**
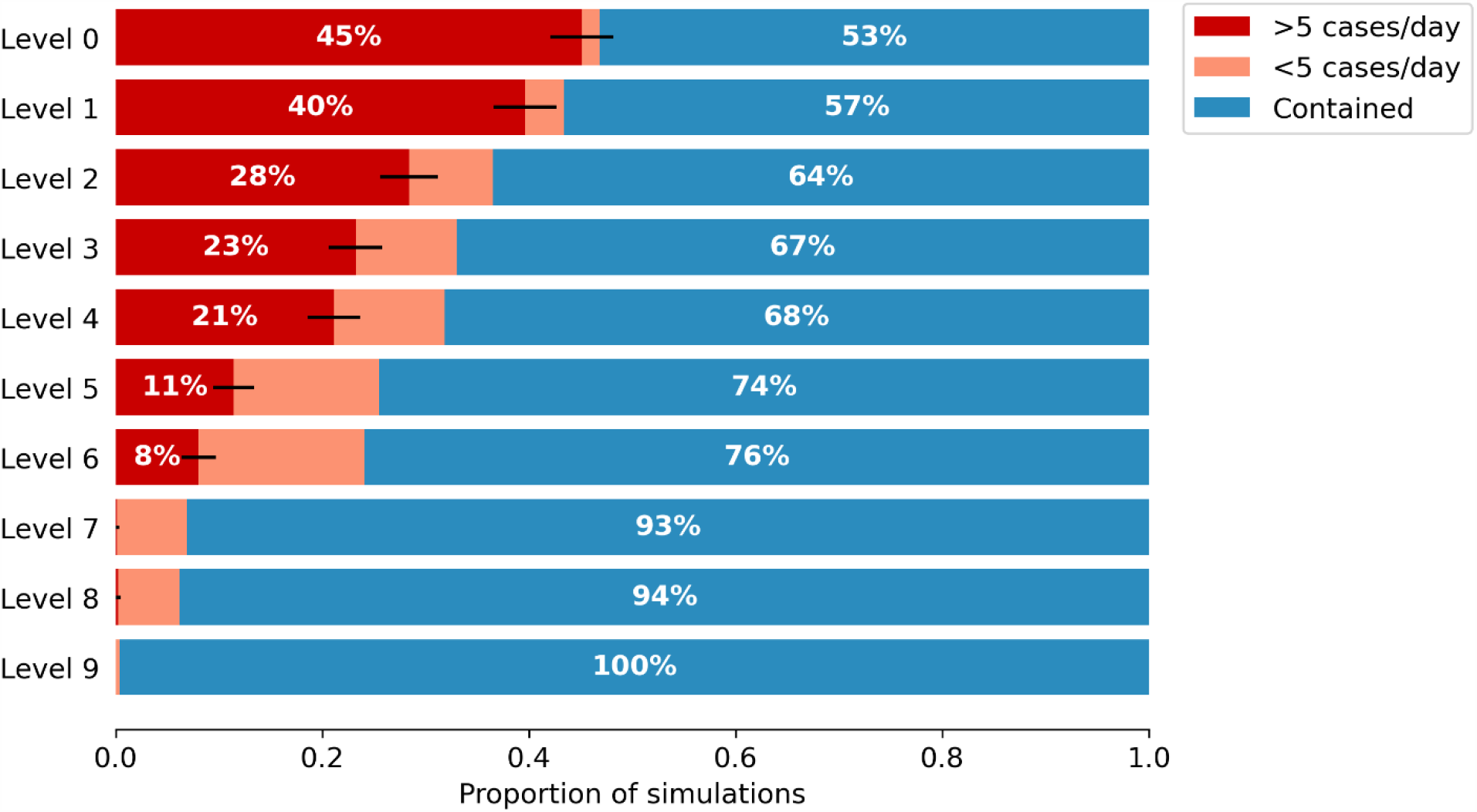
Outbreak probability. For each level of restrictions, the proportion of simulations where introducing an undiagnosed infection to a setting with zero transmission was contained (blue; defined as a 7-day average of 0 cases per day after 60 days), under control (orange; defines as a 7-day average of >0 but <5 diagnoses per day after 60 days), or led to an outbreak (red; defined as a 7-day average of >5 cases per day after 60 days). The error bars show the 95% binomial confidence interval for the 1000 simulations performed, reflecting uncertainty in the estimation of the probability for the given number of model runs.

In general, increasing the proportion of symptomatic people that seek testing is comparable or better than increasing restrictions by a single level (Figure 2). For example, if a sustainable economic option were for businesses to operate with density limits (Level 3 restrictions) then the baseline probability of an outbreak is 23%. Increasing restrictions by closing pubs and restaurants entirely (Level 4 restrictions) would only decrease the risk by 2 percentage points. However, if the testing rate was increased such that 75% of symptomatic individuals sought testing, rather than 50%, then the risk decreases by 9 percentage points. Conversely, the ‘Less testing’ scenario in which only 25% of symptomatic people seek testing has an increase in risk of 11 percentage points, highlighting the large increase in risk associated with a drop in testing.

**Figure 2.**
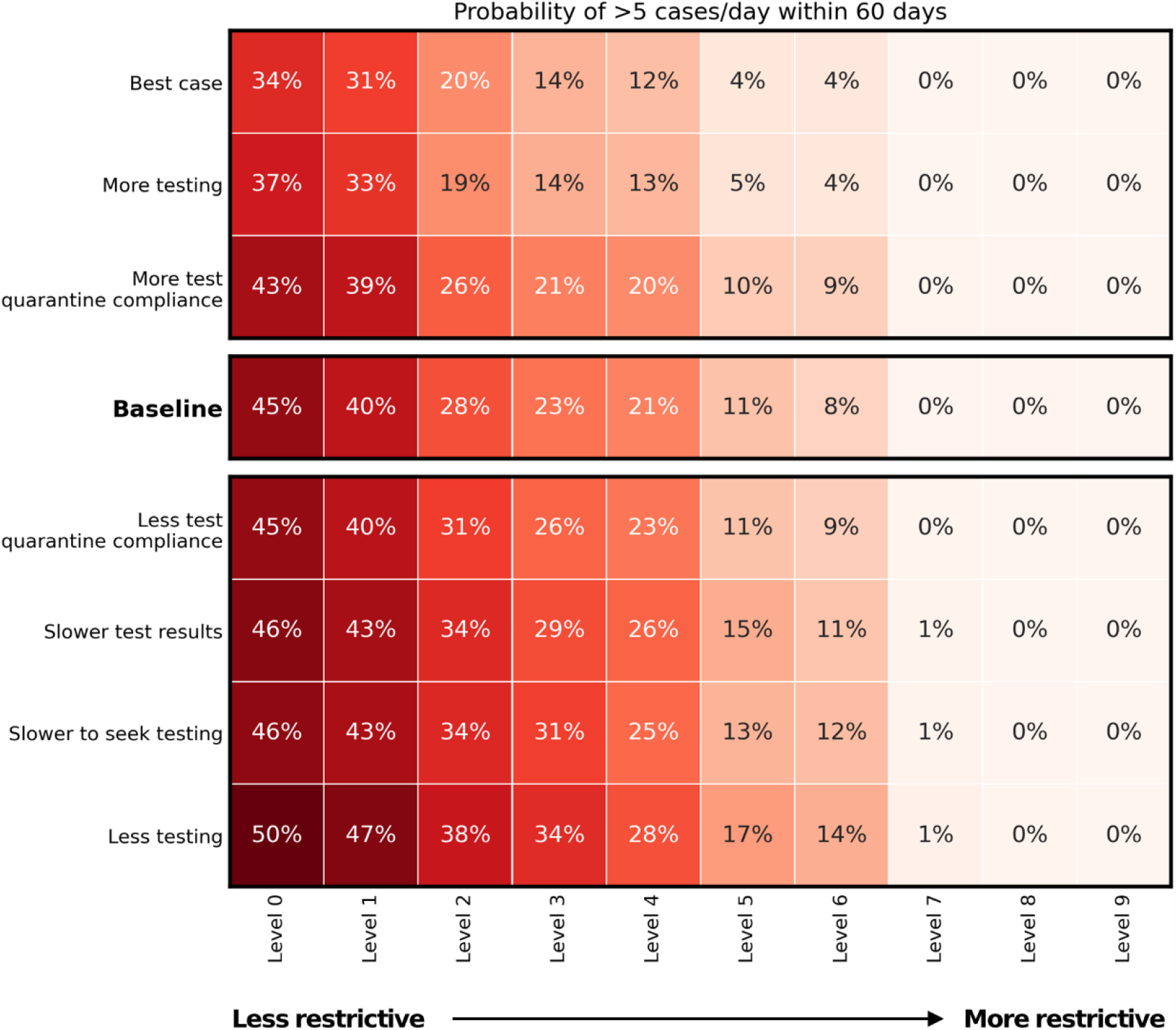
Sensitivity analysis for outbreak probability. Probability of the outbreak reaching >5 cases/day within 60 days, for each restriction level and testing/compliance combination.

Changing the proportion of people that quarantine while waiting for tests results had a minimal effect on outbreak probability. At Level 3 restrictions, full compliance only decreased risk by 2 percentage points, while 50% compliance (rather than 75%) increased risk by 3 percentage points. This relatively small effect size is likely because we have assumed that test results are returned within 24 hours, so the required quarantine period would be very small. The effect of quarantine compliance would likely be more pronounced if tests took longer to process.

### Outbreak time to first diagnosis

The time taken to diagnose the first case reflects the incubation and pre-symptomatic period for COVID-19 and is mainly driven by the testing rate. Therefore, it remained fairly consistent across all of the levels of restrictions (Figure 3). The median time to detect the outbreak was 10 days, which could decrease by up to 2 days with increased testing, or increase by up to 4 days if testing rates reduced.

**Figure 3.**
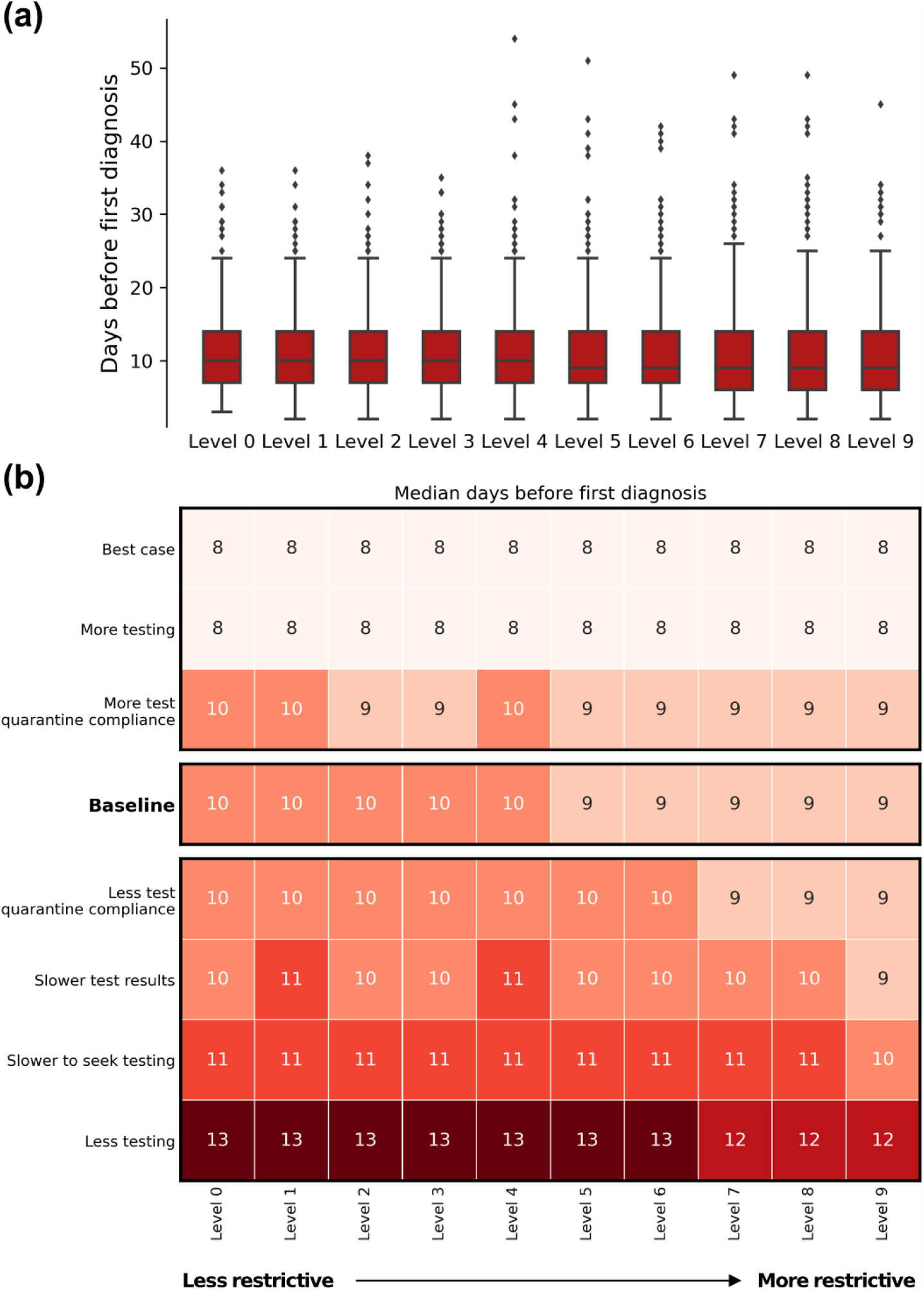
Time to first diagnosis. (a) For the baseline scenario, the distribution across the 1000 simulations sampled. (b) Median values for each restriction level and testing/compliance combination.

### Outbreak size at first diagnosis

When there were fewer restrictions and thus more rapid outbreak growth, the number of transmission events prior to the first diagnosis was higher (Figure 4). With no restrictions and 50% of symptomatic people seeking testing, there were a median of 6 community infections by the time the first diagnosis was recorded. With less testing, this increases to a median of 13 cases when the first case is diagnosed.

**Figure 4.**
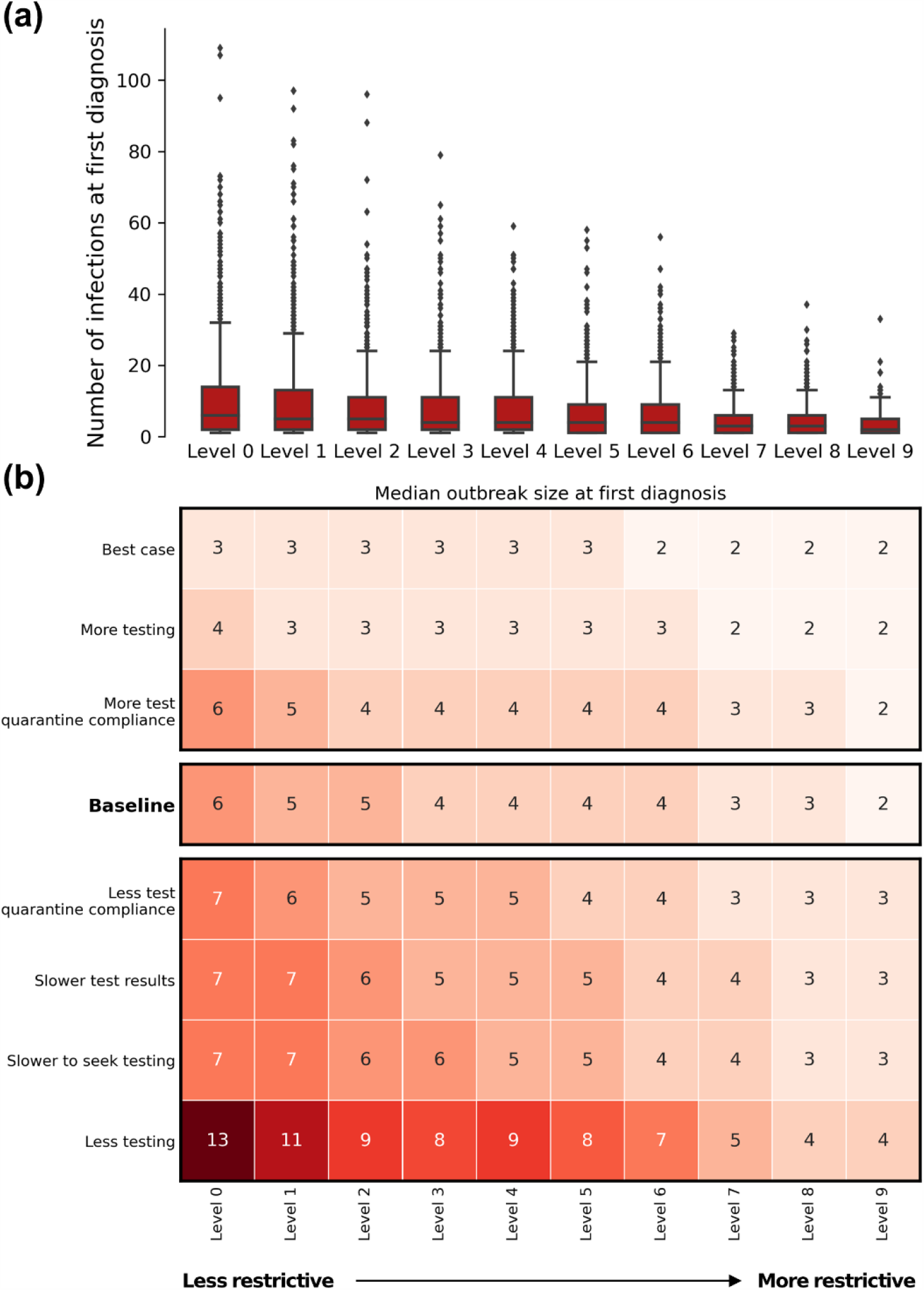
Outbreak size at first diagnosis. (a) For the baseline scenario, the distribution across the 1000 simulations sampled, (b) Median values for each restriction level and testing/compliance combination.

### Outbreak time to 5 cases/day

If the outbreak was not contained or controlled, with no restrictions the outbreak took a median of 22 days from when the first case was diagnosed to when the outbreak reached a 7-day average of 5 diagnoses per day (Figure 5). With greater restrictions in place, the growth rate was reduced. At high restriction levels, there were very few or no simulations that reached this threshold, so the distributions for Levels 8 and 9 mainly reflect the small number of samples rather than the growth rate of the outbreak.

**Figure 5.**
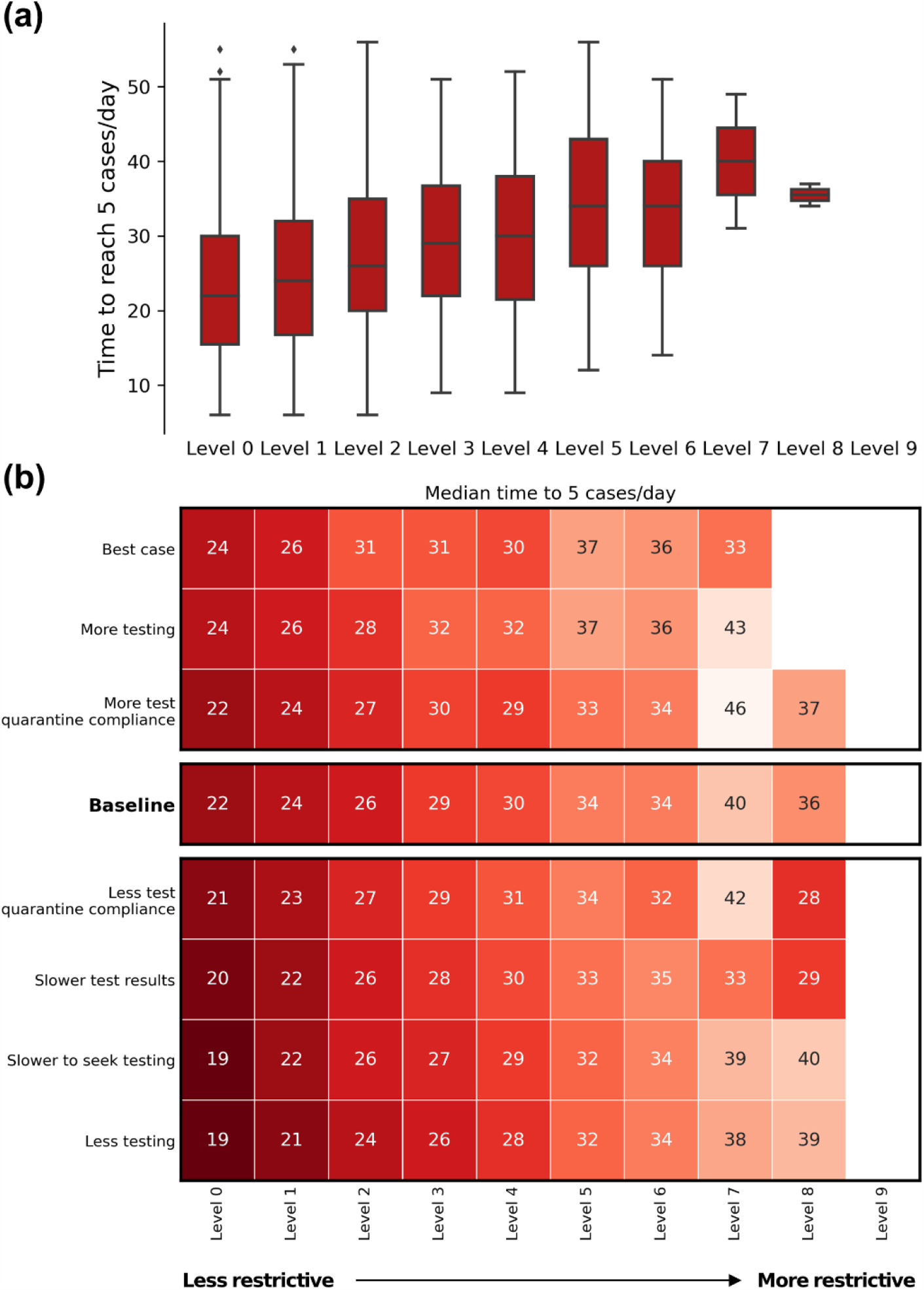
Time between the first case being diagnosed and reaching 5 cases/day. (a) For the baseline scenario, showing the distribution across the 1000 simulations sampled. (b) Median values for each restriction level and testing/compliance combination.

Overall, increases in symptomatic testing slightly increased the time to reach a 7-day average of 5 diagnoses per day, by around 2-3 days compared to baseline. This impact appears small compared to the change in outbreak risk associated with increased testing. However, increased testing means that more cases are diagnosed, which can increase the number of diagnosed cases per day even for the same total number of cases. Thus while increased testing is expected to decrease the growth rate of the epidemic due to more cases being quarantined and contact-traced, the increased number of diagnoses may partially mask this effect when looking at diagnosis-based metrics.

### Outbreak size after 60 days assuming no changes in behaviour or policy

The size of the outbreak after 60 days exhibited considerable variation, highlighting the variability in outcome depending on the specifics of who is infected and how quickly they and their contacts are identified (Figure 6). The distribution was also extremely long-tailed, with the largest possible outbreaks for each restriction level being much larger than the median or mean outcome (noting the logarithmic y-axis scale).

**Figure 6.**
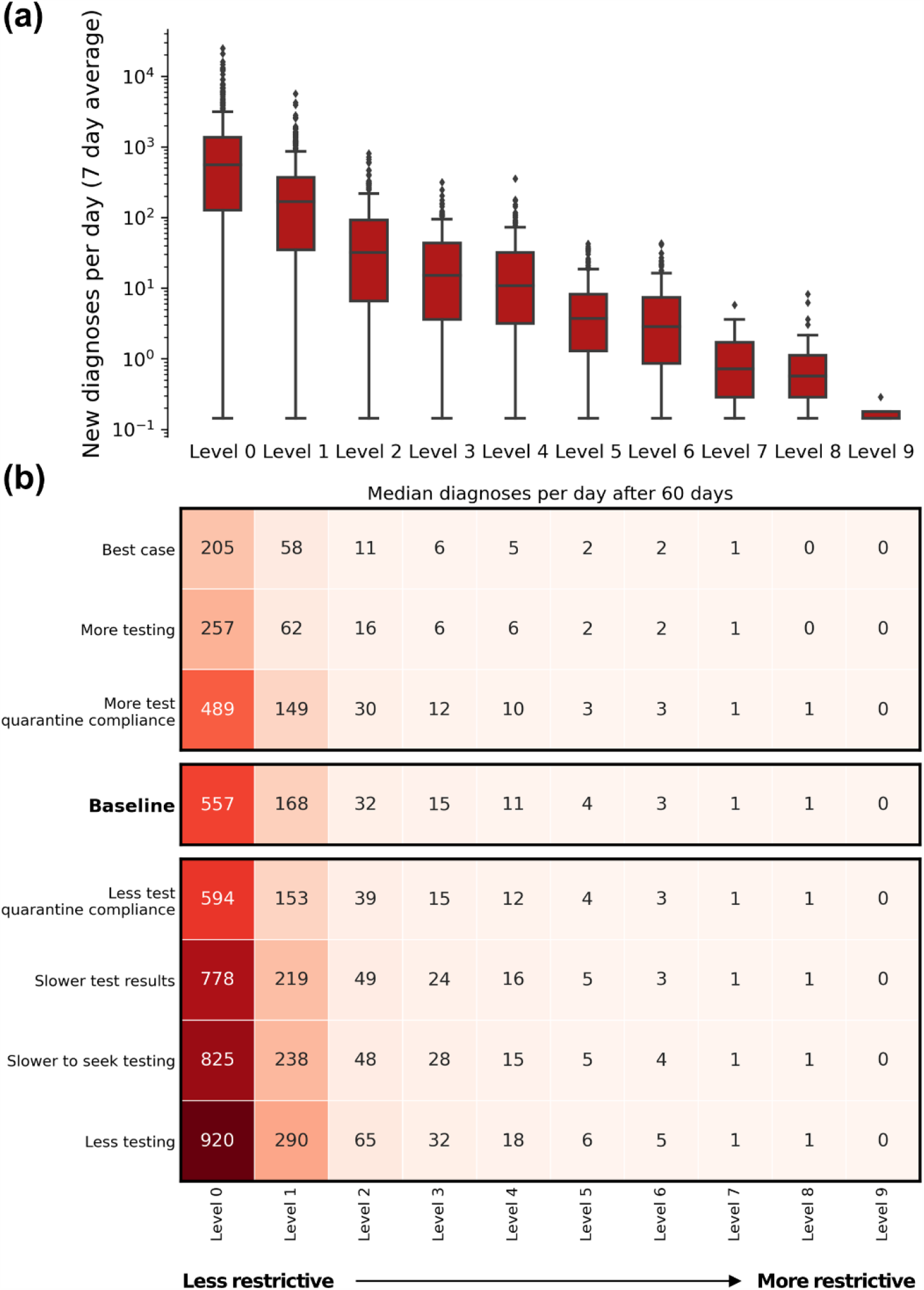
Number of new diagnoses/day (7 day average) after 60 days for each policy package, given that the outbreak was not contained (>0 diagnoses/day after 60 days, 7 day average). (a) For the baseline scenario, the distribution across the 1000 simulations sampled. (b) Median values for each restriction level and testing/compliance combination.

Even light restrictions (Level 1 and 2) resulted in much smaller outbreaks after 60 days, with median 7-day average of 32 diagnoses per day with working from home + masks (Level 2), compared to 557 diagnoses per day with no restrictions (Level 0).

We note that the simulations in Figure 6 represent worst-case counterfactual scenarios where no policy interventions take place during the 60-day simulation window. In past Australian outbreaks, interventions such as reintroduction of restrictions and suburban testing blitzes have been implemented fairly quickly (with daily diagnoses cases <30 cases/day). Similar responses would likely take place in future outbreaks, and these would change the trajectory of the outbreak compared to Figure 6.

## Discussion

We used an agent-based model to investigate possible trajectories of a newly-seeded undiagnosed COVID-19 infection in the context of no community transmission in Victoria, Australia. Our results suggest that with no restrictions in place, there is a high probability that a single introduced case could trigger an outbreak. After a seed infection, it took a median 10 days (IQR=7) to detect the first case, at which point there were estimated to be a median 6 cases (IQR=12) already in the community. In 45% of simulations an outbreak with a 7-day average of >5 diagnoses/day occurred. The results also suggest that light restrictions or behavioural changes could considerably reduce the risk of an outbreak. Working from home, working from home + masks, and working from home + masks + density limits on venues reduced the outbreak risk to 40%, 28% and 23% respectively.

These results suggest that relaxing restrictions even with no cases in the community may carry a high level of risk because a single incursion event could trigger an outbreak, and such incursion events are reasonably likely even with strict border controls. Travel-related incursions have already been observed multiple times, in Vietnam, New Zealand, and in the Australian states of Victoria, New South Wales, and South Australia. A number of studies have examined options for quarantine protocols and border controls (13-18). A recent analysis suggests that under current Australian 14-day quarantine protocols for international arrivals and based on the prevalence of COVID-19 among arrivals to Australia, there is estimated to be approximately 2 to 6 infectious days in the community per 10,000 arrivals due to the imperfect test sensitivity and possibility of >14 day incubation period of COVID-19 (17). This means that any plans to increase international arrivals from high prevalence settings, even with a 14-day quarantine, must be considered in the context of local measures to maintain a constant level of outbreak risk within a setting. An integrated risk analysis is needed to inform debates on international travel, particularly regarding how to repatriate citizens or promote economic activity (e.g. international students).

We also found that maintaining light restrictions, even when there is no community transmission, has ongoing benefits. For example, by maintaining a work-from-home policy and mandatory masks, the outbreak probability decreased to 28%, a decrease of 17 percentage points, with further decreases seen if bars and restaurants also had density limits. Moreover, maintaining light restrictions substantially reduced the growth rate of the epidemic if an outbreak did occur, providing more time to implement a response.

Maintaining or increasing symptomatic testing was another highly effective way to reduce outbreak risk, often comparable with adding a level of restrictions. For example, promoting the importance of testing such that 75% of those with symptoms test resulted in a larger decrease in outbreak risk with Level 2 restrictions than closing bars and restaurants (Level 4 restrictions). Conversely if the symptomatic testing rate were to decrease to 25% then outbreak risk increased considerably, by up to 10 percentage points. These results highlight the importance of fast, widespread testing. However, maintaining high levels of testing and timely testing (e.g. testing soon after the first symptoms) will be difficult in the context of limited or no community transmission. If several months have passed without a detected case, people are likely to assume that their symptoms are not caused by COVID-19 and may not test or delay testing to see if their symptoms are mild and/or naturally resolve. While it is true that in a setting with zero transmission, symptoms consistent with COVID-19 are unlikely to be caused by COVID-19 working to maintain high levels of symptomatic testing is critical. It increases the chance of detecting a new outbreak early, at a point when a small number of transmissions greatly influences the future trajectory of the outbreak.

Interestingly, there was only a small difference in outbreak risk depending on compliance with requirements to quarantine while waiting for test results. This is likely because in the scenarios we modelled, the number of infectious days in the community averted is limited because the test turnaround time is only one day. In the context of near-zero community transmission, the majority of people being tested are uninfected, and thus whether they quarantine or not does not make a difference to transmission. As a consequence, our results suggest it is worth exploring whether in a zero transmission setting there is a net benefit of removing the need to quarantine after testing, to increase symptomatic testing rates and detect new outbreaks more quickly. Then, if a new community case is detected, quarantine following testing could be reintroduced carte-blanche or for contacts of the case or within the relevant geographical area.

Our study builds on early work examining outbreak trajectories (18-20), incorporating extensive data from the Victorian second wave that was not previously available, and including detailed contact layers, testing, and contact tracing. In March 2020, Hellewell et al. (19) estimated that with 20 initial cases and 60% of contacts being traced, less than 50% of outbreaks would be controlled. Huamaní (20) examined new outbreaks in Peru in April 2020, and estimated that there was an 80% chance that a single infection could be contained, but that this probability decreases quickly as the number of cases increases, with only a 20% chance of containment with 10 initial cases. Our findings based on more recent data are somewhat more pessimistic, with comparable outbreak probabilities even with just one initial infection, likely driven by improved estimates of the proportion of cases that are asymptomatic, as well as the more detailed transmission dynamics in our model.

While recent studies tend to focus on containment and management of large epidemics such as those in European or American settings, analyses of new outbreaks in settings with zero community transmission remain relevant for a significant number of countries that have achieved epidemic control. As vaccines against SARS-CoV-2 become available in 2021, understanding how outbreak risk changes depending on vaccine properties and coverage will be critical for informing border controls and outbreak response plans for these settings. The approaches used in this study can be readily extended to incorporate vaccines, and we plan to investigate a range of potential vaccine rollout scenarios in future work.

## Limitations

The findings presented in this study are derived from an individual-based simulation model, Covasim. The model parameters are based on best-available data at the time of writing, including estimates of social mixing, contact networks, adherence to policies and quarantine advice, and disease characteristics (e.g. asymptomatic cases). There are several main limitations that impact the reported likelihood of an outbreak, including:

- It was assumed that after an extended period of low cases, 50% of symptomatic people would seek testing, which may be an overestimate (and hence the outbreak risk higher than estimated). However, we note that the symptomatic test rate here applies to people with COVID symptoms, and the testing rates for people with symptoms such as anosmia or fever is likely to be substantially higher than for people with mild respiratory symptoms only. Similarly, if the proportion of cases that are asymptomatic is different, the effectiveness of symptomatic testing in the community would also change accordingly.
- It was assumed that if a mandatory masks policy was in effect, compliance would be similar to at the peak of the epidemic wave in August 2020 in Melbourne. However, it is likely that after an extended period of no community transmission, mask compliance would be somewhat lower. The overall effect of masks depends on both mask efficacy and compliance, and we note that our estimates for mask efficacy are conservative.
- An outbreak was defined reaching a 7-day average of >5 new diagnoses per day within 60 days. If a different threshold, time frame, or metric (e.g. infections rather than diagnoses) were used, then the probability reported would change, although this would not substantially impact the qualitative nature of the results.
- We presented some results indicating the likely outcome after 60 days under a worst-case assumption that there is no change in official policy, testing, or other individual behavior during this time. Given that it is unlikely that a change would not occur, the results in Figure 6 should be interpreted as complementary to the results shown in Figure 1 and indicative of the potential scale of the outbreak, rather than the projections for the actual likely scale.
- In this study, we have examined scenarios where a new infection is randomly seeded into the community – for example, an interstate traveller. If an incursion occurs within a sub-population subject to different testing policies (e.g., hotel quarantine staff that are regularly tested regardless of symptoms) then the overall level of risk is likely to be lower because the case is more likely to be diagnosed early on.

## Conclusions

In a zero-transmission setting, there is an ongoing risk of a case being introduced into the community, and this risk is likely to increase as the pandemic worsens elsewhere in the world. If life ‘returns to normal’ after a period of no community transmission, there is a considerable chance that even a single introduced case could trigger an outbreak. To stop an introduced case becoming an outbreak, it is critical to detect it as early as possible. Maintaining high testing rates remains a key factor in managing the risk of future outbreaks. Further, light restrictions can substantially increase the likelihood of it remaining contained or under-control. Testing and non-pharmaceutical interventions such as masks have a large benefit with minimal impact on broader well-being and the economy. However, community support may be difficult to maintain when there are no active cases, so ongoing government and community effort/engagement will be required to ensure that societies can be as open as possible, but at the same time detect and quickly contain introduced cases to manage the risk of needing greater restrictions.

## Data Availability

Source code for the Covasim model is publicly available. A source code release for the analyses in this study is in preparation and will be available publicly.

## SUPPLEMENTARY MATERIAL

### Additional model details

The agent-based model *Covasim* models the spread of COVID-19 by simulating a collection of agents representing people. Each agent in the model is characterised by a set of demographic and disease properties:

- Demographics:
  - Age (one-year brackets)
  - Household size, and uniquely identified household members
  - Uniquely identified school contacts (for people aged 5-18)
  - Uniquely identified work contacts (for people aged 18-65)
  - Average number of daily community contacts (multiple settings / contact networks modelled, described below)
- Disease properties:
  - Infection status (susceptible, exposed, recovered or dead)
  - Whether they are infectious (no, yes)
  - An over-dispersed individual transmission factor, sampled from a negative binomial distribution with unit mean and dispersion 0.45 based on Adam, Wu (21).
  - Whether they are symptomatic (no, mild, severe, critical; with probability of being symptomatic increasing with age, and the probability of symptoms being more severe increasing with age)
  - Diagnostic status (untested vs tested)

Transmission is modelled to occur when a susceptible individual is in contact with an infectious individual through one of their contact networks. The probability of transmission per contact is calibrated to match the epidemic dynamics observed and is weighted according to whether the infectious individual has symptoms, and the type of contact (e.g. household contacts are more likely to result in transmission than community contacts). Transmission dynamics depend on the structure of these contact networks, which are randomly generated but statistically resemble the specific setting being modelled. The layers included are described below, and the model parameters values are provided for each layer that was included.

### Household contact network: household size and age structure

The household contact network was set up by explicitly modelling households. The households size distribution for Australia [5] was scaled to the number required for the number of agents in the simulation. Each person in the model was uniquely allocated to a household. To assign ages, a single person was selected from each household as an index, whose age was randomly sampled from the distribution of ages of the Household Reference Person Indicator in the 2016 Census for Greater Melbourne (22). The age of additional household members were then assigned according to Australian age-specific household contact estimates from Prem et al. (23), by drawing the age of the remaining members from a probability distribution based on the row corresponding to the age of the index member.

### School and work contact networks

The school contact network was set up by explicitly modelling classrooms. Classroom sizes were drawn randomly from a Poisson distribution with mean 21 (24). People in the model aged 5-17 years were assigned to classrooms with people their same age. Each classroom had one randomly selected adult (>21 years) assigned to it as a teacher. The result was that the school contact network was approximated as a collection of disjoint, completely connected clusters (i.e. classrooms).

Transmission in schools is influenced by age-specific disease susceptibility, and the age-specific probability of being symptomatic, which influences symptomatic testing interventions. In the model, people under 14 years have an odds ratio of 0.34 for acquiring infection relative to adults (25), and we use Victorian data to determine age-specific probability of being symptomatic, based on the percentage of positive contacts of confirmed cases who were symptomatic when they were tested. For this analysis it was additionally assumed that transmission risks in schools would be reduced by 50% relative to pre-COVID-19 based on the implementation of “COVID-Safe” plans following the second wave.

Similarly, a work contact network was created as a collection of disjoint, completely connected clusters of people aged 18-65. The mean size of each cluster was equal to the estimated average number of daily work contacts. Some workplaces are associated with a higher risk of infection, including healthcare settings, meat processing facilities, construction, warehousing and distribution, and are classified by the Department of Health and Human Services as high risk (26). In the model, we classified 15% of workplaces as high risk, based on labour force data from the Australian Bureau of Statistics (27). High risk workplaces were assigned a higher transmission probability, are less likely to be closed by restrictions (as many of these workplaces correspond to essential services.

### Additional contact networks

An arbitrary number of additional networks can be added, but for this analysis we considered those most likely to be subject to policy change. Each network layer required inputs for: the proportion of the population who undertake these activities; the average number of contacts per day associated with these activities; the risk of transmission relative to a household contact (scaled to account for (in)frequency of some activities such as pubs/bars once per week); relevant age range; type of network structure (random, cluster [as per schools/workplaces]); and effectiveness of quarantine and contact tracing interventions.

### Parameter values for each contact network

Table S1 shows the parameters that define each contact network in the model. Unless otherwise noted, parameters are derived in (5) from a mix of published and grey literature and a Delphi parameter estimation process. The columns of Table S1 refer to:

- **Mean contacts:** The average number of contacts per person in each network. Each person in the model has their individual number of contacts draw at random from a Poisson distribution with these values as the mean. For the social network layer, a negative binomial distribution was used with dispersion parameter 2 to account for a longer tail to the distribution.
- **Transmission probability:** The transmission probability per contact is expressed relative to household contacts, and reflects the risk of transmission depending on behaviour. For example, a casual contact in a public park is less likely to result in a transmission event compared to a contact on public transport.
- **Quarantine effect:** If a person is quarantined, the transmission probability is reduced by this factor. For example, an individual on quarantine at home would likely not work or use public transport, but they may still maintain their household contacts.
- **Population proportion:** Each network will only include a subset of the population e.g. every person has a household, but not every person regularly uses public transport.
- **Lower age/upper age:** Each network will only include agents whose age is within this range.
- **Clustered:** Here, we refer to a clustered network as one that consists of small groups people who are all connected to each other (e.g. classrooms), and where contacts do not change over time. This is compared to non-clustered networks, where contacts are randomly allocated. Non-clustered networks can either remain constant over time (e.g. social network) or have new contacts sampled each day (e.g. public transport).
- **Contact tracing probability –** the probability that each contact can be notified in order to quarantine

**Table S1:** Parameters for each of the networks in the model.

| Layer | Mean contacts | Transmission probability (relative to households) | Quarantine effect | Population proportion | Lower age | Upper age | Clustered | ^Contact tracing probability |
| --- | --- | --- | --- | --- | --- | --- | --- | --- |
| Household | 4 | 1 | 1 | 1 | 0 | 110 | Y | 1.00 |
| Aged care | 12 | 0.600 | 0.2 | 0.07 | 65 | 110 | Y | 0.95 |
| Schools | 21 | 0.124<sup>#</sup> | 0.01 | 1 | 5 | 18 | Y | 0.95 |
| Low risk work | 5 | 0.282 | 0.1 | 1 | 18 | 65 | Y | 0.95 |
| High risk work | 5 | 0.847 | 0.1 | 1 | 18 | 65 | Y | 0.95 |
| Church | 20 | 0.043 | 0.01 | 0.11 | 0 | 110 | Y | 0.5 |
| Community sport | 30 | 0.071 | 0 | 0.34 | 4 | 30 | Y | 0.5 |
| Childcare | 20 | 0.274 | 0.01 | 0.545 | 1 | 6 | Y | 0.95 |
| Community | 1 | 0.100 | 0.2 | 1 | 0 | 110 | N | 0.1 |
| Social | 6 | 0.124 | 0.5 | 1 | 15 | 110 | N | 0.5 |
| Entertainment | 25 | 0.008 | 0 | 0.3 | 15 | 110 | N | 0.5 |
| Cafes/Restaurants | 8* | 0.043 | 0 | 0.6 | 18 | 110 | N | 0.5 |
| Pub/bar | 8* | 0.057 | 0 | 0.4 | 18 | 110 | N | 0.5 |
| Transport | 25 | 0.164 | 0.01 | 0.114 | 15 | 110 | N | 0.1 |
| Public parks | 10 | 0.028 | 0 | 0.6 | 0 | 110 | N | 0.1 |

<sup>^</sup> Values are estimated or assumed by the authors. They do not represent data from, or the views of, the Victorian Department of Health and Human Services. The two estimates represent before and after assumed improvements in tracing systems, including the implementation of QR scanning systems in venues, media reports of locations of confirmed cases.
\*Based on proposed indoor size limits of <10 in the roadmap
<sup>#</sup>Includes a 50% reduction from pre-COVID levels based on additional public health interventions

### Testing and contact tracing

From 27th August onwards the Australian government has reported for each state the percentage of cases notifications within 24 hours of the test, and the percentage of close contacts notified within 48 hours of the positive test result (6). Recent estimates (17th September) suggest that in Victoria 100% of cases are notified in 24 hours of testing, and 99% of close contacts were notified within 48 hours of the positive test. In reality, the notification time and contact tracing time will be distributions (with these estimates suggesting that 24 hours and 48 hours are the tail ends, respectively), however the model is parametrised so that all tests and contact traces are completed at exactly the same time, and so single values are estimated as inputs. We therefore assumed that all tests are returned exactly 24 hours after they are taken, and all contacts take exactly 24 hours to be traced (the model uses daily time steps so this was selected as more appropriate than the reported tail at 48 hours (6), or than assuming no delay).

Contact tracing was modelled by selecting individuals diagnosed each day, up to a maximum of 250 people each day representing an (unvalidated) estimate of contact tracing capacity in Victoria. For each person selected, their contacts were quarantined for 14 days with a network-specific probability of being detected (Table S1), reflecting differences in the level of difficult in identifying contacts in that network. The contact tracing capacity does not apply to household contacts, which are assumed to be directly notified by newly diagnosed individuals. The limited contact tracing capacity only affects outbreaks that have grown large enough to exceed the tracing capacity – this was the case during the Victorian second wave, but most of the results in this study concern small outbreaks that are well below the tracing capacity. Only the model calibration and results for low restriction levels in Figure 6 are expected to depend on the tracing capacity.

We also assumed 25% coverage of the *COVIDSafe* app with 24-hour tracing time.

### Model calibration

The model was calibrated to the outbreak in Victoria over the June-September period, and the associated policy changes and interventions that were implemented over that period (Table S2).

**Table S2:**
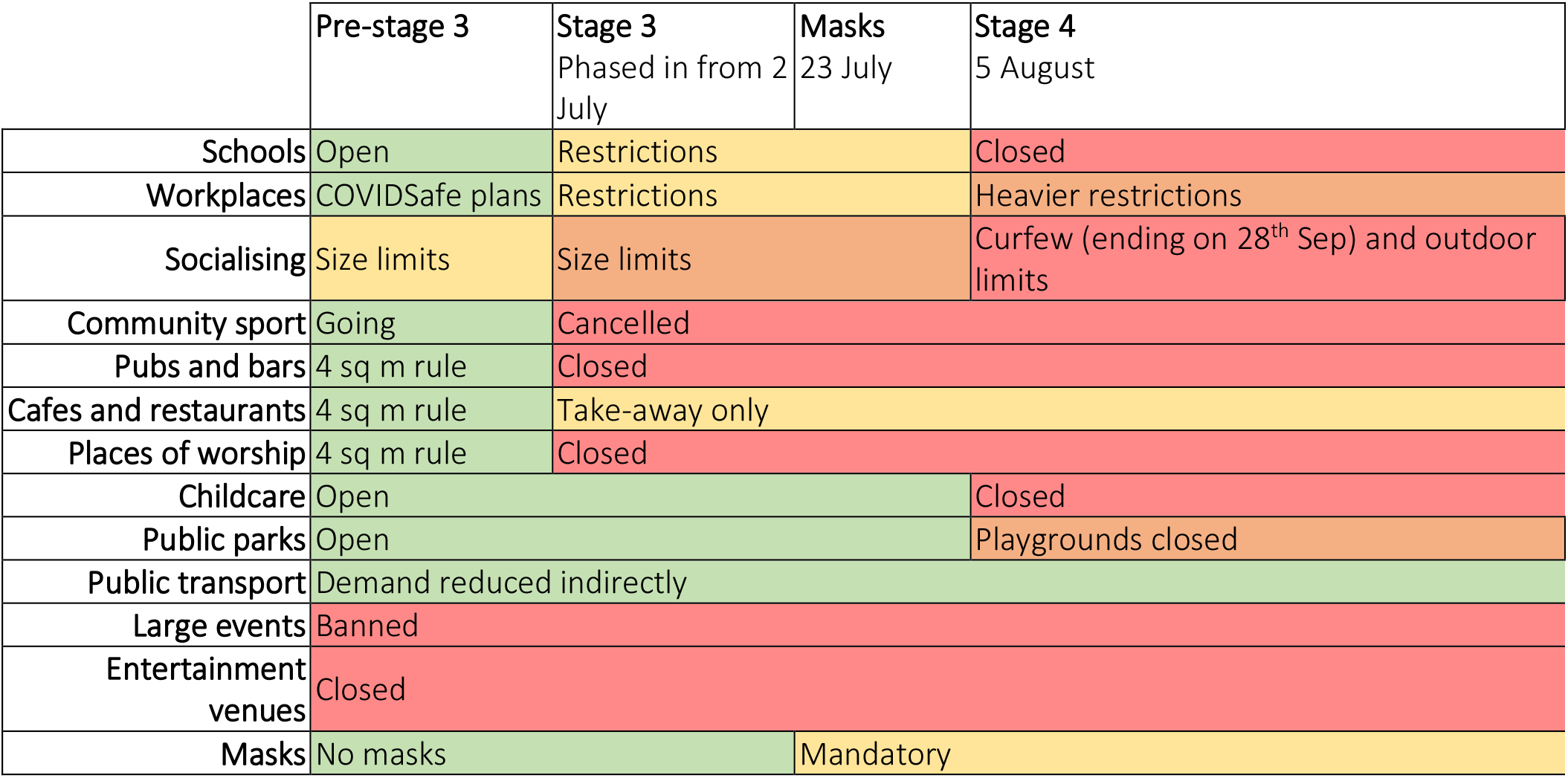
Policy changes included in the model calibration process.

Testing was modelled by assigning a per day test probability to symptomatic and asymptomatic people that was fitted as part of calibration. We assumed some improvements over time, such that there were different inputs for testing and contact tracing for June-July and August-December, with the exact day the improvements occurred calibrated to fit the epidemic trajectory. We assumed that test results took 48 hours (exactly) to be processed initially and then 24 hours (exactly) after improvement.

Overall, the transmission probability per contact governs the rate of epidemic growth, and the testing parameters affect the daily diagnoses as well as the proportion of cases that go undiagnosed. We assume that the proportion of undiagnosed cases is reflected in the number of diagnoses relative to the number of hospitalizations, as severe cases are assumed to present at hospital regardless of whether they have been tested or not. Thus, we used data on the number daily diagnoses and number of hospitalizations to enable simultaneous calibration of the testing and transmission parameters.

For the calibration shown in Figure S1, the model was initialised with a population of 100,000 agents. Due to uncertainties in the date and transmission dynamics of the original incursion events leading to the second wave, we initialized the simulation with 250 seed cases, corresponding to a point after the outbreak was already established. The overall transmission risk per contact (which multiplies the transmission probabilities in Table S1 for each layer) was varied such that when combined with inputs for the number of tests conducted over time and changes in contacts resulting from policy changes (e.g. community sports being cancelled and restaurants, cafes being take-away only when Stage 3 restrictions were introduced), the distribution of model outcomes was centred near the actual epidemic trajectory.

When calibrating, we fit the model transmission parameters under the assumption that the observed epidemic wave in June/July was the most likely outcome, which occurred in all simulations. In reality, it is possible that the second wave was an unlikely/unlucky outcome, or alternatively, that it could have been worse and was in fact a relatively lucky outcome, depending on the networks of seed cases and their contacts, as well as the overall transmission parameter. Therefore, we sampled over a set of initializations and transmission parameters, and only retained those runs where the seed/transmission parameter combination produced a projection that sufficiently matched the data – we considered the model to be a suitable fit if it was within 10% of the cumulative diagnosed cases each day. Figure S1 shows examples of the simulation runs used to estimate parameters for this study. To avoid overly penalizing mismatches in the initial stage of the outbreak, we start accumulating the cumulative case count after the 30 days, hence the model output in Figure S1(a) is offset accordingly. We note that the variability permitted in the cumulative case counts is dominated by how high the peak of the second wave is, and as the epidemic declines, the variability in new diagnoses per day by mid-September is somewhat smaller. Overall, approximately 700 of the 10000 proposed initializations were accepted. Many initializations were rejected because they diverged from the actual second wave early on, when case numbers are relatively low and the outcomes of each individual case therefore have a significant impact on the trajectory of the outbreak.

The distribution of transmission probability parameter values for the accepted initializations is shown in Figure S2.

**Figure S1:**
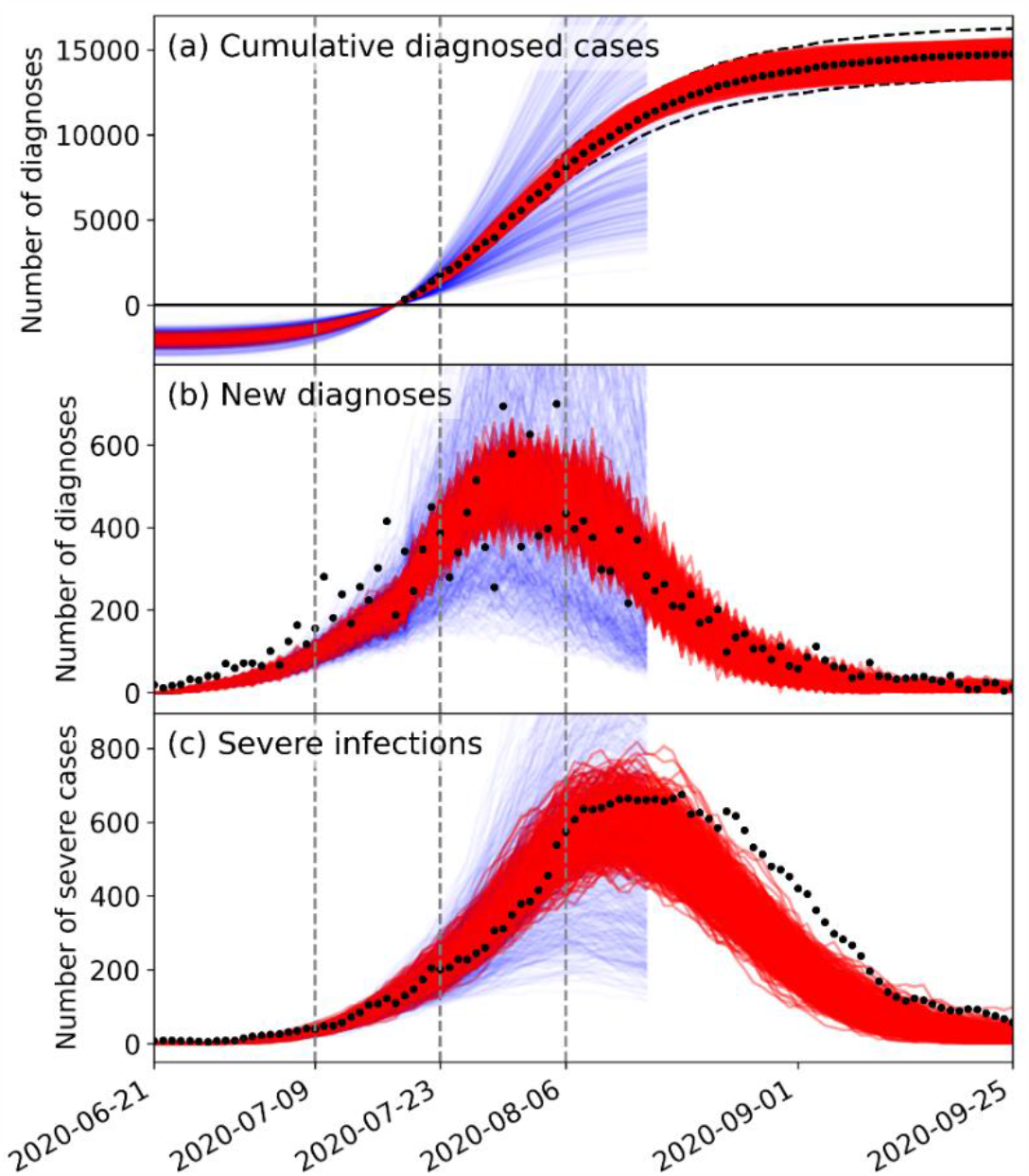
Model calibration to second wave in Victoria from June-September 2020. Vertical lines indicate when Stage 3 lockdowns took effect (9^th^ July), masks were made mandatory (23^rd^ July) and Stage 4 lockdowns took effect (6^th^ Aug). Severe infections in the model represent infections requiring hospitalisation, and the corresponding data are for reported hospitalisations. Red lines indicate simulation runs that were accepted and used to obtain the baseline beta parameter distribution used in this study; blue lines show a representative sample of simulations that were rejected.

**Figure S2:**
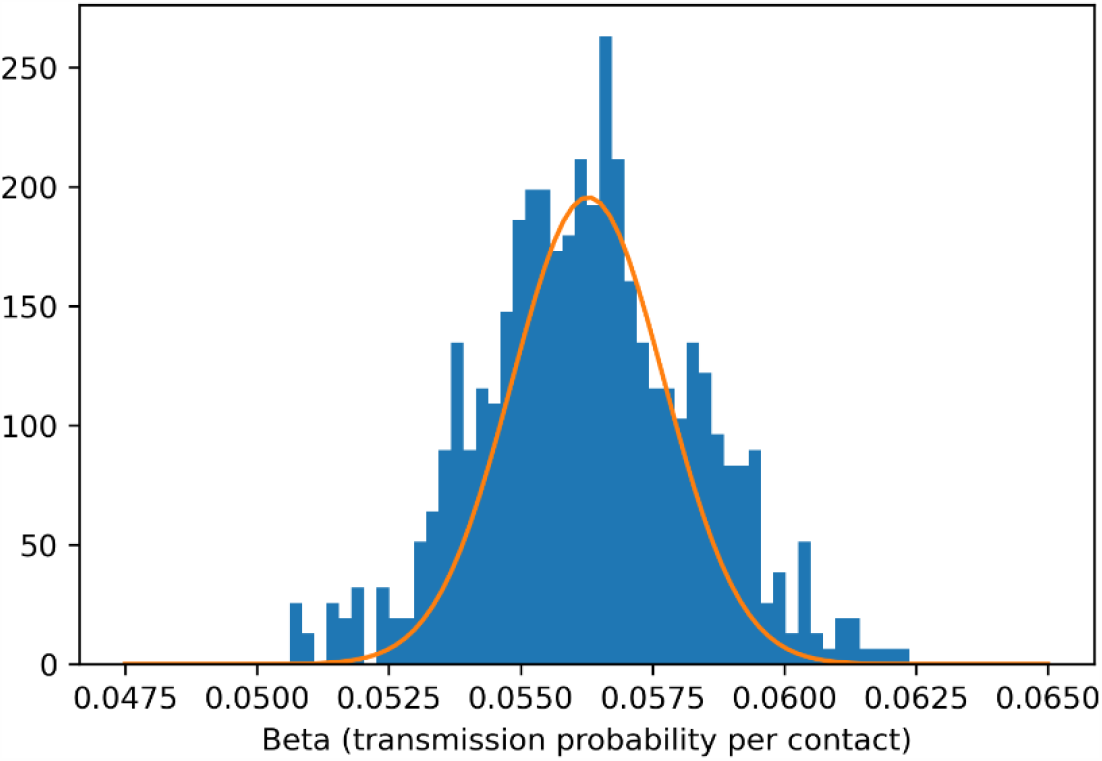
Distribution of baseline beta values for accepted calibration runs.

### Disease prognosis

**Table S3:** Age-specific susceptibility, disease progression and mortality risks.

| Age bracket | Relative susceptibility* | Prob[severe]# | Prob[critical] ## | Prob[death] ### |
| --- | --- | --- | --- | --- |
| 0-5 | 0.34 | 0.00004 | 0.0004 | 0.00002 |
| 6-12 | 0.34 | 0.00004 | 0.0004 | 0.00002 |
| 13-15 | 0.34 | 0.0004 | 0.00011 | 0.00006 |
| 16-19 | 1 | 0.0004 | 0.00011 | 0.00006 |
| 20-29 | 1 | 0.011 | 0.0005 | 0.0003 |
| 30-39 | 1 | 0.034 | 0.00123 | 0.0008 |
| 40-49 | 1 | 0.043 | 0.00214 | 0.0015 |
| 50-59 | 1 | 0.082 | 0.008 | 0.006 |
| 60-69 | 1 | 0.118 | 0.0275 | 0.022 |
| 70-79 | 1.24 | 0.166 | 0.06 | 0.051 |
| 80+ | 1.47 | 0.184 | 0.10333 | 0.093 |

\*Zhang et al. (25) found children <14 had 34% less susceptibility to adults, and people >65 years had 47% increased susceptibility

### Policies

The effect of each policy is detailed below summarized from (5), showing the impact on the transmission probability per contact, and/or the number of contacts in the network. Policies that reduce the number of contacts in the network better preserve the clustering associated – for example, the ‘Work from home’ policy reduces the number of workplace contacts to model the same people working from home every day.

#### Large events cancelled

- Large event transmission reduced by 100%

#### Entertainment venues closed

- Entertainment transmission reduced by 100%

#### Cafes/restaurants open with 4sqm physical distancing

- Cafes/restaurants transmission reduced by 50%

#### Pubs/bars open with 4sqm physical distancing

- Pubs/bars transmission reduced by 50%

#### Churches/places of worship open with 4sqm physical distancing

- Church/places of worship transmission reduced by 60%

#### Work from home where possible

- Household transmission increased by 10%
- Work transmission reduced by 36%
- Additional community transmission reduced by 33%
- Public transport transmission reduced by 33%

#### Outdoor gatherings limited to <10 people

- Additional community transmission reduced by 20%
- Entertainment transmission reduced by 100%
- Public transport transmission reduced by 50%
- Public parks transmission reduced by 40%

#### Stage 3, Melbourne and Mitchell Shire, additional impacts

- Household transmission increased by 10%
- School transmission decreases by 85%
- Community sport transmission reduced by 85%
- Cafes/restaurants transmission reduced by 85%

#### Community sports cancelled

- Community sport transmission reduced by 100%

#### Cafes/restaurants takeaway only

- Cafes/restaurants transmission reduced by 100%

#### Pubs/bars takeaway closed

- Pubs/bars transmission reduced by 100%

#### Churches and places of worship closed

- Church/places of worship transmission reduced by 100%

#### Aged care improvements

- Aged care transmission reduced by 50%

#### Mandatory masks

- Work transmission reduced by 30%
- Additional community transmission reduced by 25%
- Church/places of worship transmission reduced by 25%
- Entertainment transmission reduced by 30%
- Cafes/restaurants transmission reduced by 10%
- Pubs/bars transmission reduced by 10%
- Public transport transmission reduced by 30%
- Public parks transmission reduced by 25%
- Large event transmission reduced by 30%
- Social gatherings transmission reduced by 25%
- Aged care transmission reduced by 30%
- Schools: 0% (assumed not mandatory in these projections)

#### Small social gatherings banned

- Stage 3: social contact transmission reduced by 67%
- Stage 4: social contact transmission reduced by 90%

#### Childcare closed

- Childcare transmission reduced by 100%

#### Schools closed

- School transmission reduced by 100%

#### Mobility restrictions

- Public transport transmission reduced by 80%
- General community transmission reduced by 70%

#### Stage 4 work restrictions

- Low risk work transmission reduced by 90%
- High risk work transmission reduced by 40%

#### 50% reduction in transmissibility in Schools

- School transmission reduced by 50%

#### Outdoor gatherings limited to 50 people

- Public transport transmission reduced by 20%

## Notes

### Competing Interest Statement

The authors have declared no competing interest.

### Funding Statement

NHMRC, Burnet Institute, Bill and Melinda Gates Foundation through the Global Good Fund.

## References

1. Victorian Department of Health and Human Services. Victorian coronavirus (COVID-19) data. 2020 [Available from: https://www.dhhs.vic.gov.au/victorian-coronavirus-covid-19-data.

2. New Zealand Ministry of Health. COVID-19: Source of cases 2020 [Available from: https://www.health.govt.nz/our-work/diseases-and-conditions/covid-19-novel-coronavirus/covid-19-data-and-statistics/covid-19-source-cases.

3. Kerr C, Stuart RM, Mistry D, Abeysuriya RG, Hart G, Rosenfeld K, et al. Covasim: an agent-based model of COVID-19 dynamics and interventions. medRxiv doi:101101/2020051020097469v1.2020.

4. Institute for Disease Modeling. Covasim model. https://github.com/InstituteforDiseaseModeling/covasim.

5. Scott N, Palmer A, Delport D, Abeysuriya R, Stuart R, Kerr CC, et al. Modelling the impact of reducing control measures on the COVID-19 pandemic in a low transmission setting. Med J Aust. 2020; Accepted for publication. https://www.mja.com.au/journal/2020/modelling-impact-reducing-control-measures-covid-19-pandemic-low-transmission-setting.

6. Australian Government Department of Health. Coronavirus (COVID-19) common operating picture. https://www.health.gov.au/resources/publications/coronavirus-covid-19-common-operating-picture.

7. Abeysuriya R, Delport D, Hellard M, Scott N. Estimating risks associated with early reopening in Victoria. Policy brief. Available from: https://www.burnet.edu.au/projects/467_covasim_modelling_covid_19. mSep 2020.

8. IHME. COVID-19: What’s New for June 25, 2020. http://www.healthdata.org/sites/default/files/files/Projects/COVID/Estimation_update_062520.pdf; 2020.

9. Liang M, Gao L, Cheng C, Zhou Q, Uy JP, Heiner K, et al. Efficacy of face mask in preventing respiratory virus transmission: a systematic review and meta-analysis. Travel Medicine and Infectious Disease. 2020:101751.

10. Chu DK, Akl EA, Duda S, Solo K, Yaacoub S, Schünemann HJ, et al. Physical distancing, face masks, and eye protection to prevent person-to-person transmission of SARS-CoV-2 and COVID-19: a systematic review and meta-analysis. The Lancet. 2020.

11. New Zealand Government. COVID-19 Alert System 2020 [Available from: https://covid19.govt.nz/alert-system/.

12. UK Department of Health and Social Care. Local restriction tiers: what you need to know 2020 [Available from: https://www.gov.uk/guidance/local-restriction-tiers-what-you-need-to-know.

13. Clifford S, Quilty BJ, Russell TW, Liu Y, Chan Y-WD, Pearson CAB, et al. Strategies to reduce the risk of SARS-CoV-2 re-introduction from international travellers. 2020.

14. Wells CR, Townsend JP, Pandey A, Moghadas SM, Krieger G, Singer B, et al. Optimal COVID- 19 quarantine and testing strategies. 2020.

15. Ashcroft P, Lehtinen S, Angst DC, Low N, Bonhoeffer S. Quantifying the impact of quarantine duration on COVID-19 transmission. 2020.

16. Day M. Covid-19: Eight day quarantine is as good as 14 for returning travellers, study finds. BMJ. 2020:m3047.

17. Sacks-Davis R, Cross W, Tidhar T, Palmer A, Heath K, Scott N, et al. traQ Study: Transparet Risk Assessment of Quaranting. Final Report. Available from: https://burnet.edu.au/system/asset/file/4361/Final_Report_10November2020_Final.pdf. 2020.

18. Adekunle A, Meehan M, Rojas-Alvarez D, Trauer J, Mcbryde E. Delaying the COVID-19 epidemic in Australia: evaluating the effectiveness of international travel bans. Australian and New Zealand Journal of Public Health. 2020;44(4):257–9.

19. Hellewell J, Abbott S, Gimma A, Bosse NI, Jarvis CI, Russell TW, et al. Feasibility of controlling COVID-19 outbreaks by isolation of cases and contacts. The Lancet Global Health. 2020;8(4):e488–e96.

20. Huamaní C, Timaná-Ruiz R, Pinedo J, Pérez J, Vásquez L. Condiciones estimadas para controlar la pandemia de COVID-19 en escenarios de pre y poscuarentena en el Perú. Revista Peruana de Medicina Experimental y Salud Pública. 2020;37(2):195–202.

21. Adam DC, Wu P, Wong JY, Lau EHY, Tsang TK, Cauchemez S, et al. Clustering and superspreading potential of SARS-CoV-2 infections in Hong Kong. Nature Medicine. 2020;26(11):1714–9.

22. Australian Bureau of Statistics (ABS). Australian Bureau of Statistics 2016, Census of Population and Housing, TableBuilder. Findings based on use of ABS TableBuilder data. https://www.abs.gov.au/ausstats/abs@.nsf/web+pages/Citing+ABS+Sources#TableBuilderi. 2020.

23. Prem K, Cook AR, Jit M. Projecting social contact matrices in 152 countries using contact surveys and demographic data. PLoS computational biology. 2017;13(9):e1005697.

24. https://www.study.vic.gov.au/en/study-in-victoria/victoria's-school-system/Pages/default.aspx.

25. Zhang J, Litvinova M, Liang Y, Wang Y, Wang W, Zhao S, et al. Changes in contact patterns shape the dynamics of the COVID-19 outbreak in China. Science. 2020;368(6498):1481–6.

26. Victorian Department of Health and Human Services. High risk workplaces. https://www.dhhs.vic.gov.au/workplace-obligations-covid-19#how-do-i-know-if-my-businesses-is-a-high-risk-workplace.

27. Australian Bureau of Statistics (ABS). Census data; labour force. https://guest.censusdata.abs.gov.au/webapi/jsf/tableView/tableView.xhtml. 2016.

28. Verity R, Okell LC, Dorigatti I, Winskill P, Whittaker C, Imai N, et al. Estimates of the severity of coronavirus disease 2019: a model-based analysis. The Lancet Infectious Diseases. 2020;20(6):669–77.

29. Centers for Disease Control and Prevention COVID-19 Response Team. Severe outcomes among patients with coronavirus disease 2019 (COVID-19)—United States, February 12-March 16, 2020. MMWR Morb Mortal Wkly Rep. 2020;69(12):343–6.

30. Ferguson N, Laydon D, Nedjati Gilani G, Imai N, Ainslie K, Baguelin M, et al. Report 9: Impact of non-pharmaceutical interventions (NPIs) to reduce COVID19 mortality and healthcare demand. 2020.

